# Health economic impact of early versus delayed treatment of herpes simplex virus encephalitis in the UK

**DOI:** 10.1101/2024.02.14.24302706

**Authors:** Sylviane Defres, Patricia Navvuga, Hayley Hardwick, Ava Easton, Benedict D Michael, Rachel Kneen, Michael J Griffiths, Antonieta Medina-Lara, Tom Solomon, ENCEPH UK study group

## Abstract

**Objective:** Thanks to the introduction of recent national guidelines for treating herpes simplex virus (HSV) encephalitis health outcomes have improved. This paper evaluates the costs and the health-related quality of life implications of these guidelines.

**Design and setting:** A sub-analysis of data from a prospective, multi-centre, observational cohort ENCEPH-UK study conducted across 29 hospitals in the UK from 2012 to 2015.

**Study participants:** Data for patients aged ≥16 years with a confirmed HSV encephalitis diagnosis admitted for treatment with aciclovir were collected at discharge, 3 and 12 months.

**Primary and secondary outcome measures:** Patient health outcomes were measured by the Glasgow outcome score (GOS), modified ranking score (mRS), and the EuroQoL; health care costs were estimated per patient at discharge from hospital and at 12 months follow-up. In addition, Quality Adjusted Life years (QALYs) were calculated from the EQ-5D utility scores. Cost-utility analysis was performed using the NHS and Social Scare perspective.

**Results:** A total of 49 patients were included, 35 treated within 48 hours “early” (median [IQR] 8.25 [3.7-20.5]) and 14 treated after 48 hours (median [IQR] 93.9 [66.7 - 100.1]). At discharge, 30 (86%) in the early treatment group had a good mRS outcome score (0–3) compared to 4 (29%) in the delayed group. EQ-5D-3L utility value at discharge was significantly higher for early treatment (0.609 vs 0.221, p<0.000). After adjusting for age and symptom duration at admission, early treatment incurred a lower average cost at discharge, £23,086 (95% CI: £15,186 to £30,987) vs £42,405 (95% CI: £25,457 to £59,354) [p<0.04]. A -£20,218 (95% CI: -£52,173 to £11,783) cost difference was observed at 12-month follow-up post discharge.

**Conclusions:** This study suggests that early treatment may be associated with better health outcomes and reduced patient healthcare costs, with a potential for savings to the NHS with faster treatment.

**Article Summary:** *Strengths and limitations of this study:*

- Admissions to acute hospitals with suspected encephalitis, using predetermined inclusion criteria were recruited across 29 hospitals in the UK within a 3-year period, giving the largest cohort of prospectively recruited HSV encephalitis cases in the UK to date.
- Precise definitions to characterise those individuals with proven HSV encephalitis were applied thus ensuring accurate diagnoses.
- Individuals were followed up systematically for 12 months after discharge for clinical, and quality of life data providing the first study to assess the effect of treatment delays on health care resources, costs and health related quality of life.
- The analysis is limited by its relatively small sample size due to it being a rare disease, and the case record forms although thorough may not capture all health care costs incurred. This is particularly so for primary care and community care contact outside of the study hospitals.

## Introduction

Herpes simplex virus (HSV) encephalitis is a rare but severe brain infection, resulting in inflammation of the brain parenchyma, which causes significant morbidity and continues to have a mortality of 10%, even when treated with antiviral drugs (1,2). In the United Kingdom (UK), approximately 1 in 250,000 to 500,000 people are newly diagnosed with HSV encephalitis annually (3). Early symptoms of HSV encephalitis include flu like symptoms and lethargy which are also common to a variety of infections while the later symptoms such as speech problems and seizures tend to mimic more common brain conditions such as stroke (4). The non-specific range of symptoms have been associated with delayed diagnosis and treatment (5).

A UK study showed that the median time to treatment for suspected HSV encephalitis was 48 (range: 2-432) hours (6). Since the publication of the clinical guidelines for the management of suspected encephalitis in 2012, more recent studies have shown a reduced median time to treatment for suspected encephalitis to around 15 hours (7, 8)). But while a number of studies have reported on the favourable (modified Rankin Scale of 0–3) clinical outcomes associated with the early treatment (≤ 48 hours from admission) of HSV encephalitis (9–15), there is limited research into the potential impact on healthcare resource utilisation and costs.

We sought to evaluate the healthcare cost and health outcome implications of the time interval from hospital admission to HSV encephalitis treatment with aciclovir. We assessed the cost per quality adjusted life year and estimated the potential cost savings to the National Health Service (NHS) from improving treatment times in the UK. For the long-term impact, we hypothesise that treating HSV encephalitis patients within 48 hours would result in better neurological outcomes and lower healthcare costs.

## Methods and analysis

### Study design and setting

A sub-analysis was conducted as part a wider prospective, multi-centre, observational cohort ENCEPH-UK study on ‘Understanding and Improving the Outcome of Encephalitis’ conducted across 29 hospitals in the UK from 2012 to 2015 (16).

### Study participants

This sub-analysis was restricted to patients with confirmed HSV encephalitis aged ≥ 16 years admitted for treatment with aciclovir 10mg/kg three times daily **(Figure 1)**. Patients were stratified into early treatment for those treated within 48 hours from admission and delayed treatment for those treated ≥ 48 hours from admission. The 48 hour cut-off was chosen based, as earlier mentioned, on the evidence of significantly better neurological outcomes and health related quality of life for patients who receive treatment within this time interval from admission compared to patients with delayed treatment (6,9).

### Data collection

Patient data were collected prospectively using the standardised Case Report Forms (CRF) as part of the wider ENCEPH-UK study (16).

### Patient demographic and clinical data

Patient demographics – age and gender, clinical data – symptom duration, Charlson comorbidity index, immunocompetent status, and Glasgow Coma Score (GCS) severity were recorded on admission. Clinical outcomes including modified Rankin Scale (mRS) where 0 to 3 represented good outcomes whilst 4-6 represented poor outcomes and Glasgow Outcome Score (GOS) on a 1 (death) to 5 (good recovery) scale, were recorded at hospital discharge and at 3 and 12months follow-up.

### Resource use data and unit costs

Resource use data were collected at hospital discharge, 3 and at 12 months follow-up. This included: (1) length of hospital stay disaggregated into general ward and intensive care unit (ICU) to accurately reflect time spent in the different wards; (2) diagnostic tests [Lumbar Puncture (LP), Electroencephalogram (EEG), Magnetic Resonance Imaging (MRI) and Computerised Tomography (CT)]; (3) hospital transfer for those patients that required specialised care; and (4) outpatient follow-up costs including follow-up diagnostics, readmission and rehabilitation. Specific unit costs for hospital stay in ICU and general neurological wards were used to accurately reflect resources used in the different wards. The location of clinic appointment during follow-up was recorded which allowed for separate costs to be applied dependent on the location/type of the appointment. Data on each patients’ location at discharge and follow-up time points were recorded, noted either as home, rehabilitation or other. An average cost per episode of neurological rehabilitation for patients with an acquired brain injury was used to derive the cost of rehabilitation. Individual patient resource use data were applied to the corresponding unit costs obtained from the NHS reference costs 2018/19, drugs and pharmaceutical electronic market information tool (eMIT), 2019 where appropriate (17,18). All costs are in Great British pounds (GBP£) and a detailed breakdown of the resources utilised and unit costs are available in **Table 1**.

**Table 1:** Resource Unit Costs.

|  | Resource Use | Unit cost | Description | Source |
| --- | --- | --- | --- | --- |
| **Imaging and Diagnostics** |  |  |  |  |
|  | MRI | £183 | Diagnostic imaging (Direct access): RD03Z- Magnetic Resonance Imaging Scan of one area, with pre and post contrast. | NHS Reference Costs 2018-19 |
|  | CT scan | £138 | Diagnostic imaging (Direct access): RD22Z- Computerised Tomography Scan of one area, with pre and post contrast. | NHS Reference Costs 2018-19 |
|  | EEG | £76 | Directly Accessed diagnostic services: Conventional EEG, EMG or Nerve Conduction Studies, 19 years and over | NHS Reference Costs 2018-19 |
| **Inpatient stay** |  |  |  |  |
|  | General bed day | £317 | AA22C-AA22G: Cerebrovascular Accident, Nervous System Infections or Encephalopathy- Weighted average non-elective inpatient excess bed days | NHS Reference Costs 2018-19 |
|  | ICU bed day | £1,041 | Adult Critical care-Neurosciences adult patients: weighted average of XC05Z-XC07Z, 0-2 organs supported | NHS Reference Costs 2018-19 |
|  | Aciclovir | £10.68 | Aciclovir 500mg/20ml solution for infusion vials / Packsize 5 | Drugs and pharmaceutical electronic market information tool (eMIT)- Department of Health and Social Care |
|  |  | £2.14 | Per vial of 500mg |  |
|  | Ambulance transfer | £247 | Ambulance: ASS02- See and treat and convey | NHS Reference Costs 2018-19 |
| **Clinic/outpatient follow up** |  |  |  |  |
|  | Hospital general clinic | £152 | Total Outpatient Attendances-service code 300 | NHS reference costs 2018-19 |
|  | Neurology clinic | £168 | Total Outpatient Attendances-service code 400 | NHS reference costs 2018-19 |
|  | Infectious diseases clinic | £234 | Total Outpatient Attendances-service code 350 | NHS reference costs 2018-19 |
|  | Physiotherapy | £49 | Total Outpatient Attendances-service code 650 | NHS reference costs 2018-19 |
|  | Occupational therapy | £65 | Total Outpatient Attendances-service code 651 | NHS reference costs 2018-19 |
|  | Speech and language therapy | £97 | Total Outpatient Attendances-service code 652 | NHS reference costs 2018-19 |
|  | Psychiatry | £85 | Total Outpatient Attendances-service code 722 | NHS reference costs 2018-19 |
|  | Neuropsychology | £168 | Total Outpatient Attendances-service code 400 | NHS reference costs 2018-19 |
|  | Neuro Rehabilitation | £220 | Rehabilitation- weighted average of rehabilitation services for brain injuries and other neurological disorders levels 1-3 (service description:Other) VC60Z and VC12Z | NHS reference costs 2018-19 |
|  | Rehabilitation | £162 | Total Outpatient Attendances-service code 314 | NHS reference costs 2018-19 |
|  | Clinical psychology | £169 | Total Outpatient Attendances-service code 656 | NHS reference costs 2018-19 |
|  | Transient ischaemic attack (TIA) | £174 | Total Outpatient Attendances-service code 329 | NHS reference costs 2018-19 |
|  | Neuro community rehabilitation team | £92 | Community health services -NCRT: Neuro community rehabilitation teams | NHS reference costs 2018-19 |
| **On-going rehabilitation** |  |  |  |  |
|  | Inpatient rehab episode | £43,053 | Average episode cost of specialist inpatient neurorehabilitation stay following acquired brain injury | Turner-Stokes et al. 2016. {Turner-Stokes, 2015 #14} |

Footnote 1: NHS - National Health Service; EMIT – Electronic Marketing Information Tool; BNF – British National Formulary PSSRU – Personal Social Services Research Unit - Unit Costs for Health and Social Care

### Health-related quality of life (HRQoL) data

HRQoL data were assessed using the EQ-5D-5L, a 5 dimension generic preference based health questionnaire that measures patient reported outcomes on a) mobility, self-care, usual activities, pain/discomfort, and depression (18). The EQ-5D-5L health states defined by the EQ-5D-5L descriptive system were converted into a single index value using the ‘crosswalk’ between the EQ-5D-3L value sets and the new EQ-5D-5L descriptive system to obtain the index value for the EQ-5D-5L value sets. The EQ-5D utility values at discharge, 3 and 12 months are presented in this analysis. The health economic outcome measure was the Quality Adjusted Life Year (QALY), a summary measure of health outcomes that captures the effect of an intervention on both the quality and length of life in a single index unit, comparable across differing diseases (17,18). The QALY was generated using the EQ-5D utility values at discharge, 3 and 12months post discharge.

### Cost utility analysis

We conducted a cost-utility analysis of early treatment compared to delayed treatment of HSV encephalitis with aciclovir on the imputed data. The differences in health outcomes (QALYs) and costs between the two treatment groups at 12 months follow-up was estimated using generalised linear models (GLMs), controlling for baseline covariates. While non-parametric methods of evaluation are typically preferred, results are median estimates which are not appropriate for the decision maker, who would prefer mean estimates (19). A GLM with a gaussian family and log link was used to predict the mean QALY at 12 months follow-up (20).

A GLM model was fitted, controlling for age, gender and symptom duration at admission and treatment, to minimise their independent effect on the resource use and costs. A simpler model with only treatment group as a covariate was also fitted, however model comparison using the Akaike Information Criterion (AIC) and the Bayesian information Criterion (BIC) showed minimal differences in AIC and BIC estimates (***Appendix 1***) and as such, the model with age and symptom duration as covariates was selected to adjust for the potential confounding bias.

The mean costs and QALYs were predicted from the GLM model. The incremental cost effectiveness ratio (ICER) – a ratio of difference in predicted mean costs to difference in predicted mean QALYs were presented with their confidence intervals. To explore the uncertainty around the ICER, a non-parametric bootstrap technique with 1000 iterations was employed on the mean cost and QALY predictions. Results were presented with the bias corrected confidence intervals on the mean estimates as well as a cost effectiveness plane (CEP).

We also projected the estimated average annual savings to the NHS, based on annual incidence rate of 0.4 per 100,000 (21), if HSV encephalitis patients received treatment within 48 hours from admission. We assumed that the proportions of patients in each time to treatment group were representative of management of HSV encephalitis across the UK to scale up costs to a national level. The analysis was undertaken from the UK NHS perspective and all costs were expressed in pounds (£) for the year 2018/19. All statistical analyses were conducted in Stata/IC 14.1 for Windows.

### Missing data

Missing data were reported in variables – GCS severity, mRS and GOS scores as well as EQ-5D utility scores. Missing values were explored to assess the type of missingness. GSC severity at admission and mRS and GOS at discharge and follow-up contained missing values that were determined to be missing at random, as their probability of being missing was independent of unobserved data. Multiple imputation using chained equations (MICE) with a predictive mean matching model was used to impute the missing data on GCS severity, mRS and GOS using 20 imputations. MICE technique allows the for the simultaneous imputation of multiple variables while the predictive mean matching model maintains the original distribution structure of the data (22, 23). The Fraction of Missing Information (FMI) test statistic of 35% validated the choice of 20 imputations as sufficient.

### Statistical analyses

Patient demographics and clinical characteristics at baseline were presented as means ± standard deviations (SD), medians with interquartile ranges [IQR], and frequencies with percentages as appropriate. Group differences were assessed using the student’s t-test or Mann–Whitney U test and the Chi-squared or Fisher’s exact test for continuous and categorical variables, respectively. Group demographic clinical outcome differences were considered significant at p ≤0.05.

## Results

### Baseline demographic and clinical Characteristics

Of 341 suspected encephalitis patients recruited, 49 had HSV encephalitis and were stratified into two groups based on the timing of their first dose of aciclovir treatment **(Table 2).** 35 (71%) patients received early while 14 (29%) patients received delayed treatment, at a median of 8.3 [IQR: 3.7 to 20.5] and 93.90 [66.70 to 100.08] hours from admission, respectively. A comparison of the early versus delayed treatment groups revealed significant differences in mean age (54.3 [SD: 16.5] vs. 65.8 [SD: 16.3]; ***P=0.031***) and median symptom duration before hospital admission of (4.0 [IQR 3-7] vs. 1 [IQR 0-5]; ***P=0.014***) days, respectively.

**Table 2:** Baseline demographic characteristics and clinical features.

| Patient Characteristics | Early Treatment (n=35) | Delayed Treatment (n=14) | p-value |
| --- | --- | --- | --- |
| Age in years, mean (SD) | 54.26 (16.46) | 65.79 (16.25) | 0.031<sup>a</sup> |
| Gender, n (%) |  |  |  |
| Female | 15 (43%) | 4 (29%) | 0.52<sup>b</sup> |
| Male | 20 (57%) | 10 (71%) |  |
| Charlson comorbidity index, n (%) |  |  |  |
| No comorbidity | 27 (77%) | 8 (57%) | 0.18<sup>b</sup> |
| Comorbidity | 8 (23%) | 6 (43%) |  |
| Immunocompetent status, n (%) |  |  |  |
| Immunocompetent | 34 (97%) | 13 (93%) | 0.49<sup>b</sup> |
| Immunocompromised | 1 (3%) | 1 (7%) |  |
| GCS severity, n (%) |  |  |  |
| Severe ($\leq 8$) | 1 (3%) | 1 (7%) | 0.58<sup>b</sup> |
| Moderate (9-12) | 6 (17%) | 3 (21%) |  |
| Mild (13-15) | 28 (80%) | 10 (71%) |  |
| Symptom duration before admission in days,<br>Median [IQR] | 4.00 (3 - 7) | 1.00 (1 - 5) | 0.014<sup>c</sup> |
| Symptom duration to treatment in days,<br>Median [IQR] | 5.00 (3 - 8) | 5.00 (5 - 8) | 0.62<sup>c</sup> |

| Clinical outcomes at discharge from hospital |  |  |  |  |
| --- | --- | --- | --- | --- |
| mRS at discharge, n (%) |  |  |  |  |
|  | Good (0-3) | 30 (86%) | 4 (29%) | 0.0002<sup>b</sup> |
|  | Poor (4-6) | 5 (14%) | 10 (71%) |  |
| GOS score at discharge, n (%) |  |  |  |  |
|  | Death | 0 (0%) | 3 (21%) | 0.0243<sup>b</sup> |
|  | Persistent vegetative | - | - |  |
|  | Severe disability | 14 (40%) | 7 (50%) |  |
|  | Moderate disability | 11 (31%) | 3 (21%) |  |
|  | Good recovery | 10 (29%) | 1 (7%) |  |
| EQ-5D utility scores at discharge Median (IQR) |  | 0.61 (0.5 – 0.8) | 0.35 (0 – 0.4) | <0.0001<sup>c</sup> |

SD – Standard Deviation; IQR – inter Quartile Range; N/n – Number; GCS – Glasgow Comma Scale
Statistical tests: <sup>a</sup>Two sample t test used for normally distributed continuous variables; <sup>b</sup>Fisher's exact used for categorical variables; <sup>c</sup>Wilcoxon rank-sum used for skewed continuous variables.

Significant differences in clinical outcomes at discharge between patient treatment groups were reported for mRS *(p<0.0002)* and GOS score at discharge *(p=0.024)*. 71% (10/14) of patients in the delayed treatment group had a poor mRS score at discharge compared to 14% (5/35)) of the early treatment group. Of the 14 patients whose treatment was delayed, 3 died while only 1 patient had good recovery at discharge. 10 of the 35 patients who received treatment within 48 hours of admission had good recovery, with no neurological impairment at discharge. Patients receiving early treatment reported a significantly higher median EQ-5D utility values at discharge compared to delayed treatment patients, 0.61 (0.5 – 0.8) vs. 0.35 (0 – 0.4): ***p<0.0001***).

The results in **Table 3** below show a significant difference in the median [IQR] length of hospital stay between the early and delayed treatment groups; 31 (22–64) versus 95 (29–157) days respectively, *(**p=0.046**)*. There was no significant difference in the duration of treatment with aciclovir between the two groups *(**p=0.13**)*

**Table 3.** Patient resource use at discharge from hospital.

| Patient resources | Early treatment (n=35) | Delayed treatment (n=14) | p-value |
| --- | --- | --- | --- |
| Hospital stay in days, Median [IQR] | 31 (22 - 64) | 95 (29 - 157) | 0.0463<sup>c</sup> |
| Days on Aciclovir, Mean (SD) | 21 (8.24) | 25 (9.07) | 0.13<sup>a</sup> |

<sup>a</sup>Two sample t test, <sup>b</sup>Fisher's exact, <sup>c</sup>Wilcoxon rank-sum.

### Unadjusted mean healthcare costs

Results in **Table 4** below show that the unadjusted mean costs incurred by the patient at discharge was lower for patients receiving early treatment £22,854 (95% CI: £14,180 to £31,528) compared to delayed treatment £42,902 (95% CI: £25,859 to £59,945). The length of stay in hospital was the main driver of initial hospital admission costs, accounting for 91% of all costs incurred by patients treated early and 95% in the delayed treatment group. The mean cost of hospital stay was higher in the delayed treatment group in both the general ward and the ICU.

**Table 4:** Unadjusted Mean costs by treatment group. Results presented as means (95% CI)

| Variable | Early Treatment<br>n=35<br>(95% CI) | Delayed Treatment<br>N=14<br>(95% CI) | Difference<br>(95% CI) | p-value |
| --- | --- | --- | --- | --- |
| General ward stay | 11,929.1<br>(9002.8 to 14,855.3) | 27,668.6<br>(16,099 to 39,238.3) | -15,739.6<br>(-27,565.4 to -3,913.8) | **0.0125** |
| ICU stay | 8,939<br>(-200.18 to 18,078.2) | 13,226.1<br>(2788.4 to 23,663.8) | -4287.1<br>(-17,684.3 to 9,110) | 0.5202 |
| **Total hospital stay** | **20,868.1**<br>**(12,223.6 to 29,512.5)** | **40,894.7**<br>**(23,937.1 to 57,852.3)** | **-20,026.7**<br>**(-38,589.8 to -1,463.5)** | **0.0358** |
| Diagnostic tests (CT, MRI, EEG) | 1,654.5<br>(1,582.75 to 1,726.34) | 1,597.7<br>(1,497.54 to 1,697.9) | 56.8<br>(-62.43 to 176.09) | 0.3377 |
| Acyclovir antiviral treatment | 287.33<br>(257.05 to 317.6) | 317.86<br>(236.01 to 399.7) | -30.53<br>(-116.36 to 55.29) | 0.4634 |
| Ambulance transfer | 44.05<br>(10.29 to 77.82) | 91.8<br>(18 to 165.57) | -47.7<br>(-127.09 to 31.64) | 0.2237 |
| **Mean initial admission per patient** | **22,853.99**<br>**(14,180.36 to 31,527.61)** | **42,902.08**<br>**(25,858.86 to 59,945.29)** | **-20,048.09**<br>**(-1396.8 to 38,699.4)** | **0.0364** |
| Variable | Early Treatment<br>n=33<br>(95% CI) | Delayed Treatment<br>N=10<br>(95% CI) | Difference<br>(95% CI) | p-value |
| Outpatient clinic visits | 501.3<br>(273.13 to 729.43) | 202<br>(17.63 to 386.17) | -299.4<br>(18.89 to 579.87) | 0.0371 |
| Re-admissions at follow-up | 655.6<br>(-427.24 to 1,738.39) | 1,227.6<br>(-1,382.51 to 3,837.71) | -572<br>(-3315.23 to 2171.18) | 0.6599 |
| Follow-up diagnostics | 104.5<br>(46.91 to 162.12) | 43.4<br>(-54.78 to 141.58) | 61<br>(-47.99 to 170.22) | 0.2540 |
| Rehabilitation costs | 10,437<br>(3793.4 to 17,080.7) | 25,832<br>(9927.6 to 41,735.9) | 15,394.7<br>(-1333.1 to 32,122.6) | 0.0683 |
| **Mean follow-up cost per patient** | **11,698.5**<br>**(5126 to 18,270.9)** | **27,305**<br>**(12,411 to 42,198)** | **15,606.2**<br>**(-159.8 to 31,372.4)** | **0.0521** |
| **Total average overall costs per patient** | **34,809.3**<br>**(20,658 to 48,960)** | **71,043**<br>**(38,587.1 to 103,498.7)** | **36,234**<br>**(1926.5 to 70,540.8)** | **0.0399** |

Patients receiving early treatment incurred lower average follow-up costs of £1,216 (95% CI: £172 to £2351) compared to £1,473 (95% CI: £1166 to £4112) for those whose treatment was delayed. Similarly, the average patient cost at 12 months was lower for patients receiving early treatment in comparison to delayed treatment patients, £24,372 (95% CI: £15,007 to £33,738) versus £45,211 (95% CI: £23,014 to 67,409), respectively.

### Adjusted mean healthcare costs

Results show a higher mean patient cost at discharge of £23,086 (95% CI: £15,186 to £30,987) vs £42,405 (95% CI: £25,457 to £59,353) for patients in the early and delayed treatment groups respectively after adjusting for age and symptom duration before admission. This resulted in a difference of -£19,319 (95% C.I: -£37,783 to -£854) (**Table 5**).

**Table 5:** Covariate adjusted mean healthcare costs.

| Variable | Early Treatment<br>n=35<br>mean (95% CI) | Delayed Treatment<br>N=14<br>mean (95% CI) | Difference | p-value |
| --- | --- | --- | --- | --- |
| Total hospital stay | 21,073<br>(13,237.5 to 28,909) | 40,591.1<br>(23,287 to 57,895) | -19,518<br>(-38,211 to -825) | **0.041** |
| Diagnostic tests (CT, MRI, EEG) | 1,655.5<br>(1,583 to 1,728) | 1,595.5<br>(1,510.2 to 1,680.8) | 59.9<br>(-57.2 to 177.2) | 0.316 |
| Acyclovir antiviral treatment | 286.26<br>(257.13 to 315.39) | 320.98<br>(251.9 to 390.06) | -34.7<br>(-109.1 to 39.7) | 0.360 |
| Ambulance transfer | 55.5<br>(16.4 to 94.7) | 68.3<br>(8.6 to 127.9) | 12.74<br>(-64.04 to 89.53) | 0.745 |
| **Mean initial admission per patient** | **23,086.28**<br>**(15,185.83 to 30,986.72)** | **42,405.32**<br>**(25,456.81 to 59,353.83)** | **-19,319.04**<br>**(-37,783.33 to -854.8)** | **0.040** |
| Outpatient clinic visits | 488.2<br>(281.9 to 694.5) | 224.7<br>(71.3 to 378.1) | 263.5<br>(13 to 513.6) | 0.039 |
| Re-admissions at follow-up | 1656.3<br>(-4,064.8 to 7,377.5) | 247.6<br>(-71.4 to 566.5) | 1,408.8<br>(-4,406 to 7,223.4) | 0.635 |
| Follow-up diagnostics | 134.1<br>(-53.9 to 322.1) | 23.4<br>(-45.7 to 92.6) | 1,10.7<br>(-1,31.8 to 353.2) | **0.371** |
| Rehabilitation | 24,201.7<br>(-42,994.7 to 91,398) | 31,491.8<br>(-40,235.8 to 10,3219.4) | -7290.1<br>(-43,508.6 to 28,928.3) | **0.693** |
| **Mean follow-up cost per patient** | **16,232**<br>**(1989 to 30,475)** | **22,802**<br>**(4584 to 41,020)** | **-6570.2**<br>**(-25,326 to 12,185.2)** | **0.492** |
| **Total average overall costs per patient** | **38,358.7**<br>**(22,469 to 54,247)** | **58,576.3**<br>**(32,141.3 to 85,011)** | **-20,217.6**<br>**(-52,173 to 11,738)** | **0.215** |

Follow-up data were available on patient’s clinical outcomes during the first 12 months post inpatient stay for 43/49 patients. Four of the six patients with data missing on follow-up appointments were in the delayed treatment group. 3 patients died prior to discharge, all of whom were in the delayed treatment group, and a further 2 during the 12-month follow-up period. After adjusting for age and symptom duration before admission, there was a difference of -£6,570 (95% CI: - £25,326 to £12,185) in mean patient follow-up costs. The average costs from admission to 12 months post-discharge for patients on early treatment were £38,359 (95% CI: 22,470 to £54,247) versus £58,576 (95% CI: 32,141 to £85,011) for those receiving delayed treatment, resulting in difference of -£20,217 (95% CI: -£52,173 to £11,738).

### Cost utility analysis

**Table 6** presents a summary of the bootstrapped estimates for mean costs and QALYs for patients in the early and delayed treatment groups. The average cost per patient was higher in the delayed treatment group [£76,071 (BC 95% CI: £58,037 - £105,743) compared to the early treatment group [£34,866 (BC 95% CI: £30,715 - £39,423)]. Similarly, patients receiving early treatment reported better health outcomes with a mean QALY at 12 months of 0.613 (BC 95% CI: 0.599 - 0.630) compared to patients receiving delayed treatment 0.492 (BC 95% CI: 0.474 - 0.515).

**Table 6:** Bootstrapped adjusted Mean costs, QALYs and ICER.

| Treatment group | Mean total costs<br>(BC 95% CI) | Mean QALYs<br>(BC 95% CI) | Incremental cost<br>(95% CI) | Incremental QALY<br>(95% CI) |
| --- | --- | --- | --- | --- |
| **Early Treatment** | £34,886<br>(£30,715 - £39,423) | 0.613<br>(0.599 - 0.630) | -£41,185<br>(-£40,891 to -£38,815) | 0.125<br>(0.121 - 0.123) |
| **Delayed Treatment** | £76,071<br>(£58,037 - £105,743) | 0.488<br>(0.464 - 0.522) |  |  |

Notes 1: BC 95% CI – Bias Corrected 95% Confidence Interval; QALYs – Quality Adjusted Life Years

**Figure 2** is the Cost Effectiveness Plane (CEP) which shows that early treatment is both less costly and more effective and therefore dominates receiving delayed treatment.

## Discussion

### Principle findings from the Study

HSV encephalitis can have devastating long-term consequences for patients, with many experiencing neurological impairments (15) and on-going health issues such as memory problems epilepsy, and behavioural changes (24). Several studies have reported on the association between time to treatment with aciclovir and the clinical outcomes of patients with HSV encephalitis (12, 15). These clinical outcomes are often measured using Glasgow Outcome Scale (GOS) or modified Rankin Scale (mRS), and a number of studies found a significant association between delayed treatment with aciclovir and poor health outcomes, with the majority of studies classifying delay as greater than either 1 or 2 days (9–14,25). Although these studies have differed in the settings, populations and some of the outcome measures they used, they all showed that there is an increased chance of poor outcome with a longer time to treatment. Findings of these studies are in alignment with our analysis which found that 71% of patients in the delayed treatment groups reported a ‘poor’ mRS score at discharge compared to only 14% in the early treatment group (significant at *p<0.0002).* Similarly, for the GOS score at discharge, good recovery was reported of only 7% of the patients whose treatment was delayed compared to 29% of patients being treated early. However, these outcome measurement tools are crude and not nuanced and detailed enough to demonstrate more subtle problems that may impact on activities of daily living and employment e.g., lack of concentration, poor memory and fatigue.

No previous study has looked at the effect of treatment delays on healthcare resource, costs and HRQoL, as well as estimating cost effectiveness of early treatment. After adjusting for confounding, the mean cost at 12 months follow-up was £34,886 (BC 95% CI: £30,715 - £39,423) and £76,071 (BC 95% CI: £58,037 - £105,743) for patients receiving early and delayed treatment, respectively. It is important to note that the length of hospital stay, the main driver of healthcare costs incurred by the patients, was somewhat significantly longer for patients in the delayed treatment group (94 [IQR: 29 - 157] days) compared to the early treatment group (31 [IQR: 22 - 58] days), *(p<0.05)*. The resulting difference in the overall mean healthcare costs between the two treatment groups was - £39,960 (BC 95% CI: -£40,891 to -£38,815). QALYs were higher in the early treatment group compared to the delayed treatment group, 0.613 (BC 95% CI: 0.599 - 0.630) and 0.488 (BC 95% CI: 0.464 - 0.522) respectively. This resulted in a cost per QALY of -£334,068 (-£339,523 to -£329,856). However, statistical significance should be interpreted with caution due to the lack of power in the analysis associated with the small sample size as well as the potential selection bias which occurs when the differences in demographic and clinical characteristics prior to treatment, have an independent effect in the patient outcomes (26). These estimated healthcare costs are likely to be an underestimation of the true costs of treating HSV encephalitis as previous studies have shown patients often suffer from long term sequelae many years after initial presentation (27,28). Therefore, it is probable that additional costs to the NHS in terms of GP attendances, further diagnostic investigations and costs associated with ongoing care may have been underreported in the study. In addition, an earlier study by our group found that the sequelae after HSV encephalitis had significant impacts on activities of daily living with patients likely to incur costs through productivity losses due to difficulties returning to work either in their prior or in a changed role; there are additional costs for family/friends who become carers (24). However, given that the perspective of our analysis was that of the payer (NHS), these wider social costs were not included in the analysis.

We also projected the potential annual cost to the NHS of treating HSV encephalitis patients in the UK by assuming that the proportions of patients in each time to treatment group are representative of management of HSV encephalitis across the UK. Scaling up costs to a national level equated to average healthcare costs of £11.7 million annually, based on an annual incidence rate of 0.4 per 100,000 (21)

The differences in average healthcare costs between those treated more promptly and those delayed highlights the potential for savings for the NHS through reducing treatment delays. Significant improvements have already taken place in the UK regarding management of HSV encephalitis over the past 20 years. Our group led the development and publication of guidelines for the management of suspected viral encephalitis (21) facilitating improvements in recognition and diagnosis of the condition to reduce delays in treatment (29). Without the availability of time-series data following the same hospitals over time, it is difficult to measure the precise extent of any improvements. However, the earliest UK study (6) reported a median time to treatment of 48 hours, with 56% of patients experiencing delays of 48 hours or more, compared to 29% in this recent study. This is still too high. The national guidelines suggest all patients should be on treatment withing 6 hours of hospital admission. National level costs of a case of HSV encephalitis pre guideline publication can be estimated by assuming the proportions of patients treated early versus delayed in the earliest study (6) are representative of UK management during that time (44% treated early, 56% delayed) and applying the average costs for each treatment group estimated from this current study, adjusted for inflation using the consumer price index with 2015 as the base year (30). Based on average costs estimated in our study this would have equated to past (pre-guidelines) annual healthcare costs of £11.7 million in the UK. If the reduction in the proportion of patients experiencing delays through improvements in management over time are reflective of the UK population as a whole, this would equate to an annual saving of £720,000.

Symptoms of HSV encephalitis, particularly in the early stages of disease, have similarities with many other viral infections. However, the disease can rapidly develop over days, making prompt treatment with aciclovir crucial. There are a number of reasons that could result in treatment delays, which were not explored in this current study. Previous studies have shown that delays in undertaking CT scans can result in delays in the initiation of aciclovir (6,11), as can a failure to recognise HSV encephalitis as an initial diagnosis, due to the non-specific nature of the illness (31). This was also highlighted in a study conducted with HSV encephalitis survivors and their families, who described the difficulties in navigating health care systems during their illness trajectories, often having to develop their own care pathways in order to obtain recognition for their symptoms/ illness (3). That study emphasised the importance of involving and listening to patients and their significant others concerning symptom recognition and changes in usual behaviour to assist with the diagnosis and reduce treatment delays. In our current study, significant differences in age between the two groups were observed *(p=0.031)*, with those with delayed treatment being older on average. Differences in age may influence whether a patient is treated/diagnosed quicker for a number of reasons. For example, it may be more difficult to recognise symptoms of encephalitis in older patients, such as confusion or dizziness, and often their symptoms are attributed to other conditions common in the elderly such as delirium or stroke. Previous literature has also shown older age to be associated with poor outcomes (12,25). The observation that those delayed were older on average highlights that further improvements are needed on the management of suspected encephalitis in elderly patients, with those already at an increased chance of poor outcome being further disadvantaged through treatment delays.

### Limitations of the analysis

There are a number of limitations to this study. Firstly, as a result of the rarity of HSV encephalitis and missing data on some patient’s time of admission due to patients being transferred from other hospitals, this was a statistically small sample size. Small sample size could potentially under power the analysis and result in inadequate treatment effect estimates (32). Secondly, as is usually the case with non-randomised studies, selection bias, which was not adequately identified and adjusted for in this analysis meant that the unadjusted comparison of mean cost estimates are biased and should be interpreted with extreme caution as well as the extrapolations. Attempting to correct for selection bias with a matching (26) technique would lead to an even smaller sample size and the potential loss of informative data points. Thirdly, relying on case report forms to record patients resource use to estimate healthcare costs may not capture all the costs incurred. This is particularly true for follow-up data where primary care and community care contact outside the study hospitals will not have been captured. In addition, as is common during studies with follow-up periods, some patients had missing data in the 3- and 12-month CRFs, further reducing the sample size. There were a number of reasons for this missing data, including withdrawal due to the burden of follow-up, particularly when experiencing significant sequalae, and returning to relatives’ homes instead of their own address due to need for carers. Follow-up costs were only captured for the first 12 months post discharge and did not include primary and community care costs, which are likely to be substantial for those HSV encephalitis patients, particularly with recent studies showing the encephalitis patients often suffer from ongoing sequelae many years after initial presentation (24, 27, 28). Future studies should examine differences in time to treatment delays on longer term follow-up costs.

### Conclusions and implications

HSV encephalitis can have devastating consequences for patients. This study has shown that those treated more promptly had significantly lower inpatient stay costs than those with delayed aciclovir treatment. This was also seen during the first-year after discharge. Alongside previous literature showing that delays in treatment results in poorer patient outcomes, the results of this study show that there are also significant costs to the NHS as a consequence of these delays. Work has been undertaken in the UK over recent years to improve the management of suspected encephalitis and encourage prompt treatment with aciclovir if the condition is suspected, which has led to reductions in average treatment times. Based on the findings of this study, reducing the number of patients experiencing delays and thus improving the management of HSV encephalitis is likely to have resulted in savings to the NHS. In addition, this study indicates the potential savings that could be made in the future with further improvements in the management of HSV encephalitis in the UK. Results of this analysis validate the importance of the encephalitis management guidelines that have boosted efforts towards timely diagnosis and instigation of treatment.

## Data Availability

All data produced in the present study are available upon reasonable request to the authors

## Acknowledgements

The authors would like to thank all the participants who gave their time to help with this study, and the research nurses and principal investigators at the hospital sites for their help with recruitment. We would also like to thank Rebecca Spencer, Mike Scully, Kieran Crabtree, Katy Smith and Chris Jecks for ensuring data collection was entered from the sites.

## ENCEPH-UK Study group lead author Tom Solomon §§1

ENCEPH-UK study group members; Ruth Backman^1^, Gus Baker^2^, Nicholas J Beeching^3,4^, Rachel Breen^5^, David Brown^6^, Chris Cheyne^7^, Enitan D Carrol^1,8^, Nicholas W S Davies^9^, Sylviane Defres^1,3,4^, Ava Easton^10^, Martin Eccles^11^, Robbie Foy^12^, Marta Garcia-Finana^7^, Julia Granerod^6^, Julia Griem^13^, Michael Griffiths^1,8^, Alison Gummery^1^, Lara Harris^13^, Helen Hickey^5^, Helen Hill^5^, Ann Jacoby^2^, Hayley Hardwick^1^, Ciara Kierans^14^, Michael Kopelman^13^, Rachel Kneen^1,8^, Gill Lancaster^15^, Michael Levin^16^, Rebecca McDonald^17^, Antonieta Medina-Lara^18^, Esse Menson^19^, Benedict Michael^1^, Natalie Martin^20^, Andrew Pennington^17^, Andrew Pollard^20^, Julie Riley^17^, Manish Sadarangani^20^ Anne Salter^21^, Kukatharmini Tharmaratnam^7^ Maria Thornton^17^, Angela Vincent^22^, Charles Warlow^23^.

1. Institute of Infection, Veterinary and Ecological Sciences, University of Liverpool, Liverpool, UK
2. Department of Clinical Neuropsychology, The Walton Centre NHS Foundation Trust, Liverpool, UK
3. Tropical and Infectious Disease Unit, Liverpool University Hospitals NHS Foundation Trust, Liverpool, UK
4. Clinical Trials Unit, Liverpool, UK
5. Clinical Sciences, Liverpool School of Tropical Medicine, Liverpool
6. UK Health Security Agency (formerly Public Health England) Colindale, London, UK
7. The Department of Biostatistics, Institute of translational medicine, University of Liverpool, Liverpool, UK
8. Alder Hey Hospital Children’s NHS Foundation Trust, Liverpool, UK
9. Department of Neurology, Chelsea and Westminster NHS Trust, London, UK
10. Encephalitis Society, Malton North Yorkshire, UK
11. Institute of Health and Society, Newcastle University, Newcastle, UK
12. Faculty of Medicine and Health, Leeds, Institute of Health Sciences, Leeds University, Leeds, UK
13. Institute of Psychiatry, Kings College London, London, UK
14. Public Health and Policy, Institute of psychology Health and Society, University of Liverpool, Liverpool, UK
15. Mathematics and Statistics, Lancaster University, Lancaster, UK
16. Paediatrics and International Child Health, Imperial College London, UK
17. Research and Development Department, The Walton Centre NHS Foundation Trust, Liverpool, UK
18. Health Economics Group, University of Exeter medical School, Exeter, UK
19. Infectious diseases and Immunology team, Evelina London Children’s Hospital, London, UK
20. Oxford Vaccine Group, University of Oxford, Oxford, UK
21. Patient representative, Encephalitis Society, Malton, North Yorkshire
22. Nuffield Department of Clinical Neurosciences, University of Oxford, Oxford, UK
23. Department of Neurosciences, Western General Hospital, University of Edinburgh, Edinburgh, UK

Hospitals sites involved in recruitment into ENCEPH UK

## Principal Investigators

Gavin Barlow^1^, Nicholas J Beeching^2^, Thomas Blanchard^3^, Richard Body^4^, Gavin Boyd^5^, Lucia Cebria-Prejan^6^, David Chadwick^7^, Richard Cooke^8^ Pamela Crawford^9^, Brendan Davies^10^, Nicholas W S Davies^11^, Sam Douthwaite^12^, Hedley Emsley^13^, Simon Goldenberg^12^, Clive Graham^14^, Steve Green^15^, Clive Hawkins^10^, Dianne Irish^16^, Kate Jeffrey^17^, Matt Jones^18^, Liza Keating^19^, Jeff Keep^20^, Susan Larkin^8^, Maria Leita^17^, Derek Macallan^21^, Jane Minton^22^, Kavya Mohandas^23^, Ed Moran^24^, David Muir^25^, Monicka Pasztor^26^, Matthew Reed^27^, Tom Solomon^28^, Philip Stanley^29^, Julian Sutton^30^, Peter Thomas^31^, Guy Thwaites^12^, John Weir^19^, Mark Zuckerman^20^.

1. Hull and East Yorkshire Hospitals NHS Trust, Hull.
2. Liverpool University Hospitals NHS Foundation Trust Liverpool
3. North Manchester General Hospital, Manchester
4. Manchester Royal Infirmary, Manchester
5. Calderdale and Huddersfield NHS Foundation Trust, Huddersfield
6. Mid Yorkshire Hospitals NHS Trust, Pindersfield
7. South Tees Hospitals NHS Foundation Trust, Middlesborough
8. Aintree University Hospital NHS Foundation Trust, Liverpool
9. York Teaching Hospital NHS Foundation Trust, York
10. University Hospitals North Midlands, Stoke on Trent
11. Chelsea and Westminster Hospital NHS Foundation Trust, London
12. Guy’s and St Thomas’ NHS Foundation Trust, London
13. Lancashire Teaching Hospitals NHS Foundation Trust, Royal Preston Hospital, Preston
14. North Cumbria University Hospitals NHS Trust, Carlisle
15. Sheffield Teaching Hospitals NHS Foundation Trust, Sheffield
16. Royal Free London NHS Foundation Trust, London
17. Oxford University Hospitals NHS Foundation Trust, John Radcliffe Hospital, Oxford
18. Salford Royal NHS Foundation trust, Salford
19. Royal Berkshire NHS Foundation Trust, Reading
20. Kings College Hospital NHS Foundation Trust, London
21. St George’s University Hospitals NHS Foundation Trust, London
22. Leeds Teaching Hospitals NHS Trust, St James’ University Hospital, Leeds
23. Wirral University Teaching Hospital NHS Foundation Trust - Arrowepark Hospital, Upton
24. University Hospitals Birmingham NHS Foundation Trust, Birmingham Heartlands Hospital, Birmingham
25. Imperial College Healthcare NHS Trust, St Mary’s Hospital, London
26. University Hospitals of Morecambe Bay NHS Foundation Trust, Lancaster
27. NHS Lothian, Royal Infirmary Edinburgh, Edinburgh
28. The Walton Centre NHS Foundation Trust, Liverpool
29. Bradford Teaching Hospitals NHS Foundation Trust, Bradford
30. University Hospital Southampton NHS Foundation Trust, Southampton
31. Milton Keynes University Hospital NHS Foundation Trust, Milton Keynes

## ENCEPH-UK contributors

Gavin Barlow, Nicholas J Beeching, Thomas Blanchard, Richard Body, Gavin Boyd, Lucia Cebria-Prejan, David Chadwick, Richard Cooke, Pamela Crawford, Brendan Davies, Nick Davies, Sam Douthwaite, Hedley Emsley, Simon Goldenberg, Clive Graham, Steve Green, Clive Hawkins, Dianne Irish, Kate Jeffrey, Matt Jones, Liza Keating, Jeff Keep, Susan Larkin, Maria Leita, Derek Macallan, Jane Minton, Kavya Mohandas, Ed Moran, David Muir, Monicka Pasztor, Matthew Reed, Tom Solomon, Philip Stanley, Julian Sutton, Peter Thomas, Guy Thwaites, John Weir, Mark Zuckerman. Ruth Backman, Gus Baker, Rachel Breen, David Brown, Chris Cheyne, Martin Eccles, Robbie Foy, Julia Granerod, Julia Griem, Alison Gummery, Lara Harris, Helen Hickey, Helen Hill, Ann Jacoby, Ciara Kierans, Michael Kopelman, Rachel Kneen, Gill Lancaster, Michael Levin, Rebecca McDonald, Esse Menson, Natalie Martin, Andrew Pennington, Andrew Pollard, Julie Riley, Manish Sadarangani, Maria Thornton.

## Footnotes

### Author contributions

SD, AML, TS devised the idea around the health economic analysis. SD wrote the protocol, coordinated with the multiple sites in the study, checked data and wrote the paper along with PN and TS. AML supported PN and SD with the economic analyses, data interpretation and drafting of the manuscript. PN performed the economic and statistical analyses. HH and SD submitted the ethics and research and development applications and coordinated the multiple sites.SD, HH AE BDM RK MJF AML and TS formed the steering committee of the study. All authors contributed to reviewed and approved the final draft of the paper.

### Patient consent

The study protocols were approved by participating sites and the National Research Ethics Service (now part of the Health Research Authority) East Midlands Nottingham 1 committee (reference 11/EM/0442). Written consent for entry into the study was obtained from patients or an accompanying relative. Standardised case record forms for clinical, laboratory and radiological data were recorded on a secure online database (OpenclinicaTM).

### Data sharing statement

No additional data available.

## References

1. Davies N. Herpes Simplex virus encephalitis [Internet]. 2019 [cited 2019 Nov 4]. Available from: https://www.encephalitis.info/herpessimplexvirusencephalitis

2. Sabah M, Mulcahy J, Zeman A. Herpes simplex encephalitis. BMJ Br Med J [Internet]. 2012 Jun 6;344:e3166. Available from: http://www.bmj.com/content/344/bmj.e3166.abstract

3. Cooper J, Kierans C, Defres S, Easton A, Kneen R, Solomon T. Diagnostic pathways as social and participatory practices: The case of herpes simplex encephalitis. PLoS One. 2016 Mar 1;11(3).

4. Easton A, Atkin K, Dowell E. Encephalitis, a service orphan: The need for more research and access to neuropsychology. Br J Neurosci Nurs [Internet]. 2006 Dec 1;2(10):488–92. Available from: 10.12968/bjnn.2006.2.10.22531

5. Hughes PS, Jackson AC. Delays in Initiation of Acyclovir Therapy in Herpes Simplex Encephalitis. Can J Neurol Sci [Internet]. 2019 [cited 2019 Nov 14];39:644–8. Available from: 10.1017/S0317167100015390

6. Bell DJ, Suckling R, Rothburn MM, Blanchard T, Stoeter D, Michael B, et al. Management of suspected herpes simplex virus encephalitis in adults in a UK teaching hospital. Clin Med J R Coll Physicians London. 2009;9(3):231–5.

7. Defres S, Tharmaratnam K, Michael BD, Ellul M, Davies NWS, Easton A, et al. (2023) Clinical predictors of encephalitis in UK adults–A multi-centre prospective observational cohort study. PLoS ONE 18(8): e0282645.

8. Backman R, Foy R, Diggle PJ, Kneen R, Easton A, Defres S, et al. (2018) A pragmatic cluster randomised controlled trial of a tailored intervention to improve the initial management of suspected encephalitis. PLoS ONE 13(12): e0202257.

9. McGrath N, Anderson NE, Croxson MC, Powell KF. Herpes simplex encephalitis treated with acyclovir: Diagnosis and long term outcome. J Neurol Neurosurg Psychiatry. 1997;63(3):321–6.

10. Dagsdóttir HM, Sigurðardóttir B, Gottfreðsson M, Kristjánsson M, Löve A, Baldvinsdóttir GE, et al. Herpes simplex encephalitis in Iceland 1987–2011. Springerplus. 2014;3(1).

11. Poissy J, Wolff M, Dewilde A, Rozenberg F, Raschilas F, Blas M, et al. Factors associated with delay to acyclovir administration in 184 patients with herpes simplex virus encephalitis. 2009; Available from: https://liverpool.idm.oclc.org/login?url=https://search.ebscohost.com/login.aspx?direct=true&db=edsbas&AN=edsbas.A9DF2DBC&site=eds-live&scope=site

12. Singh TD, Fugate JE, Hocker S, Wijdicks EFM, Aksamit Allen J. J, Rabinstein AA. Predictors of outcome in HSV encephalitis. J Neurol Off J Eur Neurol Soc [Internet]. 2016;263(2):277. Available from: https://liverpool.idm.oclc.org/login?url=https://search.ebscohost.com/login.aspx?direct=true&db=edssjs&AN=edssjs.D3F4F273&site=eds-live&scope=site

13. Tan IL, McArthur JC, Venkatesan A, Nath A. Atypical manifestations and poor outcome of herpes simplex encephalitis in the immunocompromised. Neurology. 2012 Nov 20;79(21):2125–32.

14. Khaled M, Youssof AMA, Mourad HS. Clinical and Laboratory Predictors of Outcome in Patients with Herpes Simplex Encephalitis. Egypt J Neurol Psychiatry Neurosurg [Internet]. 2014 Apr;51(2):173–80. Available from: https://liverpool.idm.oclc.org/login?url=https://search.ebscohost.com/login.aspx?direct=true&db=a9h&AN=96002955&site=eds-live&scope=site

15. Raschilas F, Wolff M, Delatour F, Chaffaut C, De Broucker T, Chevret S, et al. Outcome of and Prognostic Factors for Herpes Simplex Encephalitis in Adult Patients: Results of a Multicenter Study. Clin Infect Dis. 2002 Aug;35(3):254–60.

16. Defres S, Tharmaratnam K, Michael BD, Ellul M, Davies NWS, Easton A, Griffiths MJ, Bhokaj M, Das K, Hardwick H, Cheyne C, Kneen R, Medina-Lara A, Salter AC, Beeching NJ, Carrol E, Vincent A; ENCEPH-UK study group; Garcia-Finana M, Solomon T. Clinical predictors of encephalitis in UK adults - A multi-centre prospective observational cohort study. PLoS One. 2023 Aug 23;18(8):e0282645. doi: 10.1371/journal.pone.0282645. eCollection 2023.PMID: 37611003 Free PMC article.

17. Drugs and pharmaceutical electronic market information tool (eMIT) [Internet]. 2019 [cited 2019 Nov 1]. Available from: https://www.gov.uk/government/publications/drugs-and-pharmaceutical-electronic-market-information-emit

18. NHS trust and NHS foundation trusts. National Health Service: National Cost Collection for the NHS - Year 2018-19 [Internet]. 2019 [cited 2019 Nov 1]. Available from: https://www.england.nhs.uk/national-cost-collection/

19. Drummond M. Methods for the Economic Evaluation of Health Care Programmes. Fourth edi. Sculpher MJ, Claxton K, Stoddart GL, Torrance GW, Stoddart GL, editors. Oxford, England: Oxford University Press; 2015. 445 p.

20. Mihaylova B, Briggs A, O’Hagan A, Thompson SG. Review of statistical methods for analysing healthcare resources and costs. Health Econ. 2011 Aug;20(8):897–916

21. Solomon T, Michael BD, Smith PE, Sanderson F, Davies NWS, Hart IJ, et al. Management of suspected viral encephalitis in adults – Association of British Neurologists and British Infection Association National Guidelines. J Infect [Internet]. 2012 Apr 1 [cited 2019 Nov 4];64(4):347–73. Available from: https://www.sciencedirect.com/science/article/pii/S0163445311005639?via%3Dihub

22. Paton NI, Stöhr W, Oddershede L, Arenas-Pinto A, Walker S, Sculpher M, et al. Health economics analysis. 2016 [cited 2019 Jan 11]; Available from: https://www.ncbi.nlm.nih.gov/books/NBK350294/

23. Wulff JN, Ejlskov L. Multiple imputation by chained equations in praxis: Guidelines and review. Electron J Bus Res Methods. 2017;15(1):41–56.

24. Cooper J, Kierans C, Defres S, Easton A, Kneen R, Solomon T. Care beyond the hospital ward: understanding the socio-medical trajectory of herpes simplex virus encephalitis. BMC Health Serv Res [Internet]. 2017 Dec 12 [cited 2020 Feb 19];17(1):646. Available from: http://bmchealthservres.biomedcentral.com/articles/10.1186/s12913-017-2608-2

25. Poissy J, Champenois K, Dewilde A, Melliez H, Georges H, Senneville E, et al. Impact of Herpes simplex virus load and red blood cells in cerebrospinal fluid upon herpes simplex meningo-encephalitis outcome. BMC Infect Dis [Internet]. 2012 Dec 18 [cited 2020 Feb 19];12(1):356. Available from: https://bmcinfectdis.biomedcentral.com/articles/10.1186/1471-2334-12-356

26. Faria R, Alava MH, Manca A, Wailoo AJ, Uk WWNO. NICE DSU TECHNICAL SUPPORT DOCUMENT 17: THE USE OF OBSERVATIONAL DATA TO INFORM ESTIMATES OF TREATMENT EFFECTIVENESS IN TECHNOLOGY APPRAISAL: METHODS FOR COMPARATIVE INDIVIDUAL PATIENT DATA REPORT BY THE DECISION SUPPORT UNIT [Internet]. 2015 [cited 2020 Feb 19]. Available from: www.nicedsu.org.uk

27. Ramanuj PP, Granerød J, Davies NWS, Conti S, Brown DWG, Crowcroft NS. Quality of Life and Associated Socio-Clinical Factors after Encephalitis in Children and Adults in England: A Population-Based, Prospective Cohort Study. Bayer A, editor. PLoS One [Internet]. 2014 Jul 29 [cited 2020 Feb 19];9(7):e103496. Available from: https://dx.plos.org/10.1371/journal.pone.0103496

28. Granerod J, Davies NWS, Ramanuj PP, Easton A, Brown DWG, Thomas SL. Increased rates of sequelae post-encephalitis in individuals attending primary care practices in the United Kingdom: a population-based retrospective cohort study. J Neurol [Internet]. 2017 Feb 1 [cited 2020 Feb 19];264(2):407–15. Available from: http://www.ncbi.nlm.nih.gov/pubmed/27766471

29. Encephalitis: recent advances and challenges ahead - ACNR | Online Neurology Journal ACNR | Online Neurology Journal [Internet]. [cited 2020 Feb 19]. Available from: https://www.acnr.co.uk/2012/12/encephalitis-recent-advances-and-challenges-ahead/

30. Consumer price index (2010 = 100) - United Kingdom | Data [Internet]. [cited 2020 Mar 3]. Available from: https://data.worldbank.org/indicator/FP.CPI.TOTL?end=2019&locations=GB&name_desc=true&start=2008

31. Hughes PS, Jackson AC. Delays in initiation of acyclovir therapy in herpes simplex encephalitis. Can J Neurol Sci [Internet]. 2012 Sep [cited 2020 Feb 19];39(5):644–8. Available from: http://www.ncbi.nlm.nih.gov/pubmed/22931707

32. Pourhoseingholi MA, Vahedi M, Rahimzadeh M. Sample size calculation in medical studies. Gastroenterol Hepatol from Bed to Bench. 2013;6(1):14–7.

